# Maternal influenza vaccination during pregnancy and risk of ASD in the offspring

**DOI:** 10.1101/2025.06.21.25330046

**Authors:** Shahar Neeman, Maor Hemo, Gal Meiri, Dorit Shmueli, Idan Menashe

## Abstract

**Background:** Vaccination has been proposed as a potential risk factor for autism spectrum disorder (ASD), contributing to public hesitancy and mistrust toward immunization. Influenza vaccination during pregnancy is considered safe and effective in preventing serious maternal complications and adverse birth outcomes associated with influenza infection. However, limited research exists regarding the long-term impact of maternal influenza vaccination on offspring neurodevelopment.

**Methods:** We conducted a retrospective cohort study of all singleton live births among members of Clalit Health Services (CHS), Israel’s largest healthcare provider, between January 2016 and December 2020, with follow-up through May 2024. Offspring with developmental disorders of known genetic etiology were excluded. Maternal influenza vaccination during pregnancy (exposure) and offspring ASD diagnosis (outcome) were identified via CHS electronic medical records. The association between exposure and outcome was assessed using Cox proportional hazards models, adjusting for sociodemographic, maternal, and gestational covariates.

**Results:** Of 153,321 children included in the analysis, 39,361 (25.7%) were exposed to maternal influenza vaccination during pregnancy. A modestly increased risk of ASD was observed in the crude analysis among offspring of mothers who received influenza vaccination during pregnancy compared to those who did not (HR = 1.22, 95% CI: 1.14–1.31). However, this association was no longer evident after adjustment for sociodemographic and clinical covariates (aHR = 0.97, 95% CI: 0.91–1.05).

**Conclusions:** Maternal influenza vaccination during pregnancy was not associated with an increased risk of ASD in offspring. These results support the long-term safety of maternal influenza vaccination as a preventive healthcare measure.

## Introduction

Autism spectrum disorder (ASD) is a neurodevelopmental condition characterized by persistent challenges in social communication and repetitive behaviors. The prevalence of ASD has risen consistently over recent decades and is now estimated at 2.2% globally, with a male:female ratio of approximately 4:1.(1) While the exact causes of ASD remain unclear, maternal infections during pregnancy, especially viral infections and infections requiring hospitalization, have been suggested as possible risk factors.(2–5)

The influenza virus is associated with increased mortality and morbidity in high-risk populations, such as older adults, individuals with obesity or type 2 diabetes, immunocompromised individuals, and pregnant women.(3) Influenza infection during pregnancy poses considerable risks to both mother and fetus. Specifically, influenza infection in pregnant women may cause respiratory distress and other severe influenza-related complications that may lead to hospitalization and, for high-risk pregnant women, even to admission to intensive care units.(4–6) The risks to newborns of affected mothers include intrauterine growth restriction (IUGR) and congenital malformations.(7)

Vaccination for influenza viruses has been available for more than 80 years. (8) The inactivated influenza vaccine has been demonstrated to be safe and effective in all trimesters of pregnancy, with no evidence of increased maternal or fetal risk.(9) Consequently, the World Health Organization (WHO) recommends influenza vaccination for pregnant women as a priority group.(10) Nevertheless, public concerns remain regarding the potential neurodevelopmental outcomes, particularly ASD, of this vaccination. To date, only a few large cohort studies have investigated the association between influenza vaccination during pregnancy and a subsequent diagnosis of ASD in the offspring resulting in mixed results.(11–13) Other studies reported no significant associations between maternal influenza vaccination and adverse infant neurodevelopmental outcomes, although these studies did not specifically evaluate ASD risk.(14,15) Given the limited and mixed information about the long-term neurodevelopmental implications of gestational influenza vaccination, further research into this important area is warranted.

## Methods

### Study Setting and Population

We conducted a retrospective cohort study of all singleton live births between January 2016 and December 2020 in a population belonging to Clalit Health Services (CHS), Israel’s largest healthcare provider, serving over half of the country’s population. The study cohort drawn from mothers and neonates who were members of CHS at the time of birth included singleton births at a gestational age of at least 24 weeks that occurred in CHS-affiliated hospitals. We excluded children with known genetic syndromes associated with ASD, such as Down’s syndrome, Fragile X syndrome, Prader-Willi syndrome, Williams syndrome, tuberous sclerosis, and Turner syndrome. A flow diagram of the study cohort ascertainment is presented in **Fig. 1**. All children in the study were followed until May 2024 or until an ASD diagnosis was determined, ensuring that all children had a potential follow-up time of at least 3.5 years.

**Fig. 1.**
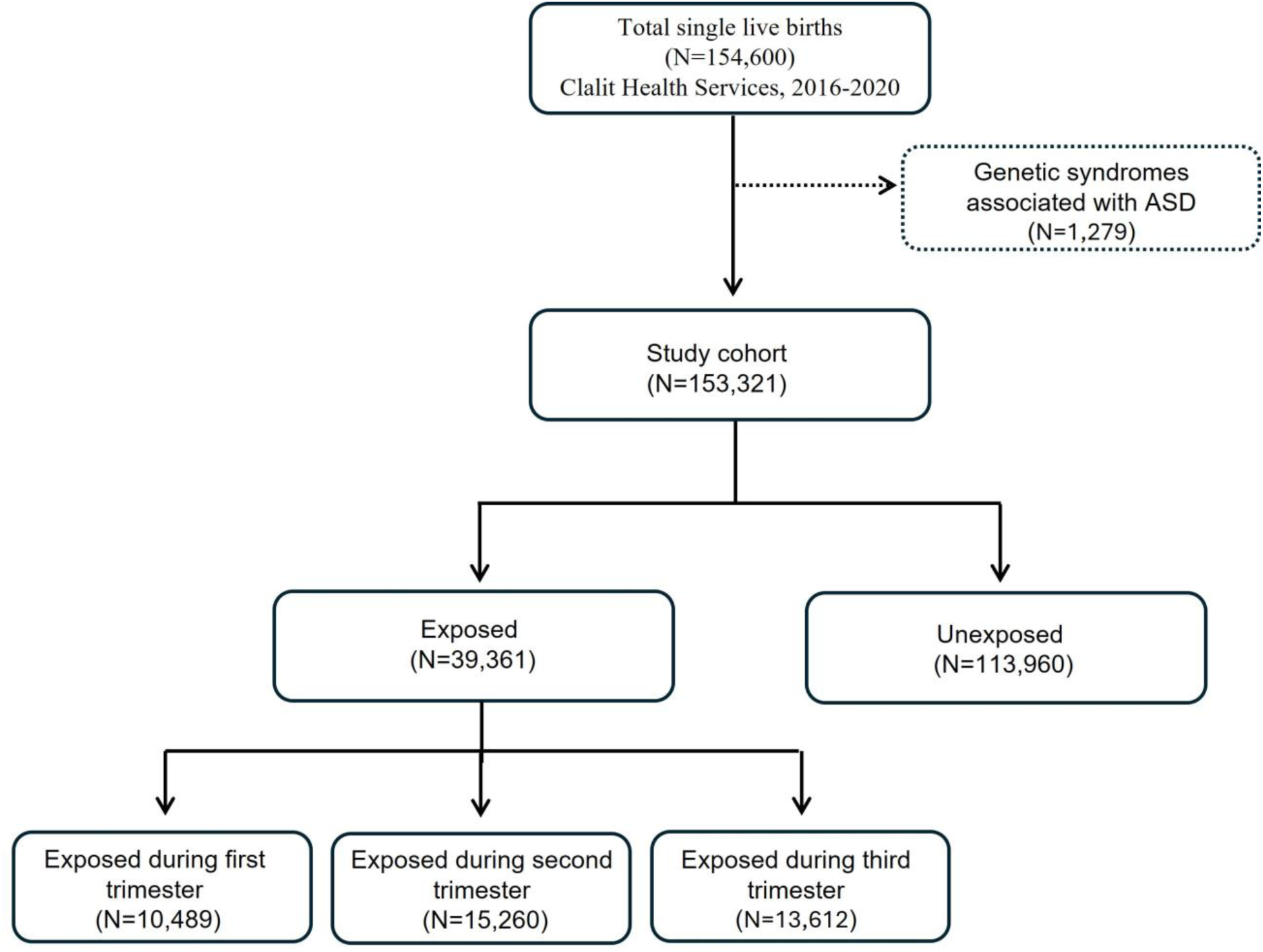
Flow Diagram of the Study Cohort Ascertainment and Vaccinated (Exposed) Groups

## Study Variables

All data for this study were obtained from CHS electronic database via the MDclone system and subsequently analyzed in the virtual desktop infrastructure (VDI) environment of CHS. The *exposure* variable was defined as maternal influenza vaccination during pregnancy, documented as the administration of at least one dose of influenza vaccine between the date of conception and the date of birth. The *outcome* variable was defined as a diagnosis of ASD as recorded in the CHS database (because documentation of an ASD diagnosis, according to DSM-5 criteria(16) is a prerequisite for state-funded benefits, including ASD-related health services, it is assumed that most children members of CHS with a formal diagnosis of ASD are documented as such in the CHS database). Additional *covariates* were extracted from CHS medical records to account for factors potentially influencing the association between maternal vaccination and ASD. These variables were selected based on their association with both the exposure and outcome variables, as reported in the literature.

Maternal age at delivery was categorized into four groups: under 25, 25–34, 35–39, and ≥40, reflecting distinct maternal and obstetric risk profiles across these age ranges. Maternal body mass index (BMI) before pregnancy was classified according to medical guidelines into underweight (<18.5 kg/m²), normal weight (18.5–24.9 kg/m²), overweight (25–29.9 kg/m²), and obese (≥30 kg/m²). Gravidity was divided into three categories: 1, 2–4, and ≥5, reflecting the clinical importance of first and high-parity pregnancies. Smoking status was classified as either smoker or non-smoker, based on information collected from medical documents. While this data provided valuable insights into smoking behaviors, it did not necessarily reflect smoking habits specifically during pregnancy. Ethnicity was grouped into three categories: Jews, Arabs, and others. The district of residence was categorized as South, North, or Center, with Jerusalem being included in the central district. Regarding maternal medical history, inflammatory bowel disease, asthma, and diabetes mellitus were recorded as either "yes" or "no." Pregnancy-related variables were also analyzed, including gestational diabetes, oligohydramnios, polyhydramnios, the use of antibiotics during pregnancy, a history of preterm labor, and documented cases of decreased fetal movements. Delivery type was classified into vaginal delivery, assisted vaginal delivery, or Cesarean section. Vaginal delivery included both normal deliveries and breech deliveries, while assisted vaginal delivery included the use of forceps or vacuum. Gestational age at birth was classified as preterm (<37 weeks) or term (≥37 weeks). Birth weight was included as a continuous variable, with mean (SD), median, minimum, and maximum values being recorded.

## Statistical Analysis

Relevant confounders for the main analysis were selected based on their association with both the exposure and outcome variables. The statistical significance of such associations was determined via univariate analyses, such as Chi-square test for the association between the exposure/outcome and other categorical variables and t-test or Wilcoxon’s rank-sum test for the association with continuous or ordinal variables, respectively. Kaplan-Meier survival curves were used to assess the cumulative incidence of ASD in the offspring of mothers vaccinated against influenza during pregnancy or not vaccinated—the exposed and unexposed groups, respectively. Cox proportional hazards regression models were employed to estimate the HR of ASD in the offspring associated with gestational influenza vaccination, while adjusting for the selected covariates. Censoring of participants in the Kaplan-Meier and Cox analyses was due to a child’s death or loss to follow-up (e.g. disenrollment from CHS). All analyses were conducted using R (version 4.2.3).

## Ethics

The research protocol was approved by the Helsinki Committee of the Soroka University Medical Center, Beer-Sheva, Israel (SOR #0244-23).

## Results

Of the 154,600 live singleton births to mothers belonging to CHS, 153,321 met the inclusion and exclusion criteria for the study cohort (**Fig. 1**). Of these births, 39,361 (25.7%) were to mothers who had received influenza vaccination during pregnancy (exposed group), and 113,960 (74.3%) were to unvaccinated mothers (unexposed group). Among the exposed group, 10,489 (26.6%) mothers had been vaccinated during the first trimester, 15,260 (38.8%), during the second trimester and 13,612 (34.6%), during the third trimester (**Fig. 1**). The median follow-up time of the study cohort was 5.9 years.

**Table 1** presents the sociodemographic, maternal and gestational characteristics for the study cohort. Most mothers in the study were Jewish (62.3%), living in either the center (43.3%) or the south (40.4%) of the country. There were significant differences between the exposed and unexposed groups in terms of sociodemographic and maternal characteristics. Specifically, for the exposed group, there was a higher percentage of Jewish mothers living in the center of the country, whereas the non-exposed group included a higher proportion of Arab mothers living in southern Israel (74.9% Jewish and 22.8% Arab making up the exposed group vs. 57.9% Jewish and 39.3% Arab for the unexposed group, respectively, and 54.0% Center and 26.1% South in the exposed group vs. 39.5% and 45.3% for the unexposed group). In addition, mothers in the exposed group were slightly older than those in the unexposed group, with a higher proportion of women aged ≥25 years (87.0% vs. 79.6%) and a higher proportion of smokers (11.3% vs. 9.0%). In terms of pregnancy-related variables, gestational comorbidities, such as gestational diabetes (8.4% vs. 6.6%) and oligohydramnios (3.3% vs. 2.8%), were more prevalent in the exposed group. Additionally, asthma (25% vs. 22.8%) and diabetes mellitus (3.3% vs. 2.0%) were slightly more prevalent in the exposed group.

**Table 1.**
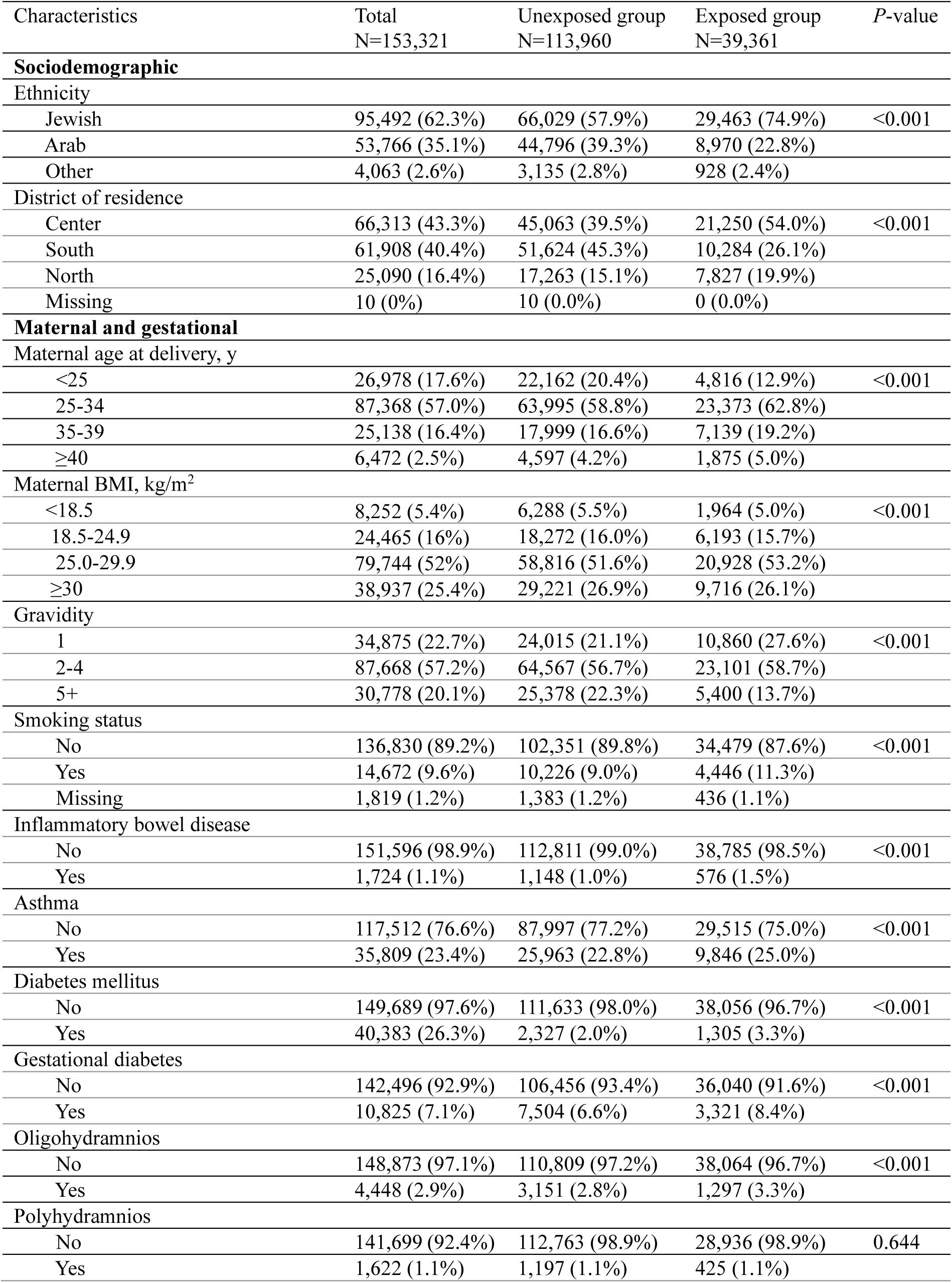
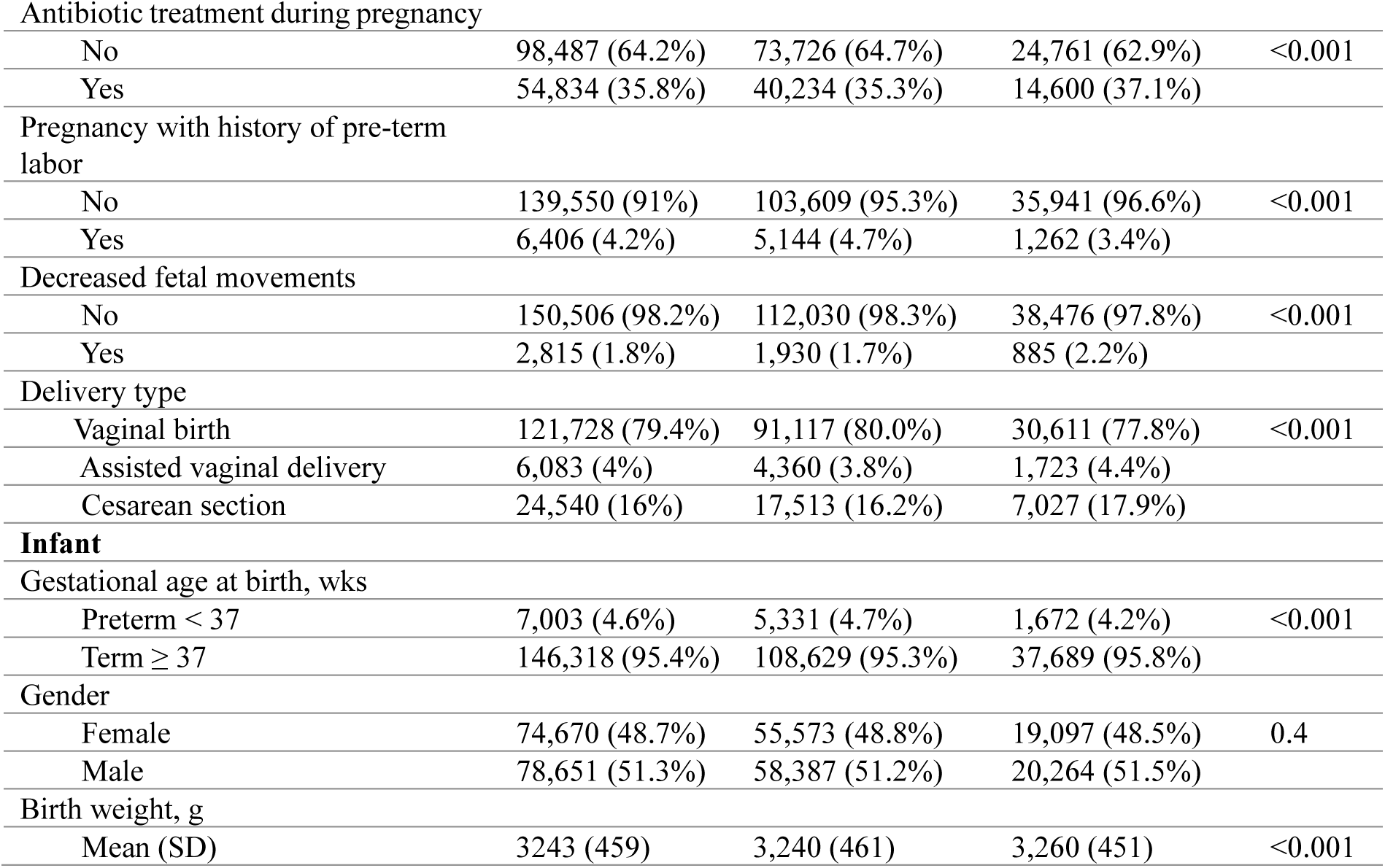
Maternal and gestational characteristics of the study cohort

We also compared maternal characteristics for children with and without ASD (**Table 2**) to identify characteristics associated with both the *exposure* and *outcome* variables, thus highlighting them as potential confounders. Here, too, large differences between the groups were seen in the ethnicity and district of residence of participants, in that the subgroup of Jewish mothers from central Israel had the highest proportion of ASD children compared to all other subgroups (82.3% vs. 61.8% for Jewish ethnicity and 57.6% vs. 42.9% for Center district). Other notable differences between mothers of ASD and non-ASD children were evident in the proportion of smokers (16.2% vs, 9.4%), maternal age (87.8% vs. 81.3% of mothers ≥25 years), Cesarean deliveries (25.4% vs. 16.4%), and rates of gestational diabetes (12% vs. 6.9%).

**Table 2.** Maternal and gestational characteristics of mothers of ASD and non-ASD children

| Characteristics | No ASD group<br>N=149,645 | ASD group<br>N=3,676 | *P*-value |
| --- | --- | --- | --- |
| **Sociodemographic** |  |  |  |
| Ethnicity |  |  |  |
| Jewish | 92,465 (61.8%) | 3,027 (82.3%) | <0.001 |
| Arab | 53,199 (35.6%) | 567 (15.4%) |  |
| Other | 3,981 (2.7%) | 82 (2.2%) |  |
| District of residence |  |  |  |
| Center | 64,196 (42.9%) | 2,117 (57.6%) | <0.001 |
| South | 60,918 (40.7%) | 990 (26.9%) |  |
| North | 24,522 (16.4%) | 568 (15.5%) |  |
| Missing | 9 (0.0%) | 1 (0.0%) |  |
| **Maternal and gestational** |  |  |  |
| Maternal age at delivery, y |  |  |  |
| <25 | 26,547 (18.6%) | 431 (12.3%) | <0.001 |
| 25-34 | 85,335 (59.9%) | 2,033 (58.0%) |  |
| 35-39 | 24,351 (17.1%) | 787 (22.5%) |  |
| >40 | 6,218 (4.4%) | 254 (7.2%) |  |
| Maternal BMI, kg/m<sup>2</sup> |  |  |  |
| <18.5 | 8,028 (5.4%) | 224 (6.1%) | <0.001 |
| 18.5-24.9 | 23,661 (15.8%) | 804 (21.9%) |  |
| 25.0-29.9 | 78,062 (52.2%) | 1,682 (45.8%) |  |
| ≥30 | 39,894 (26.7%) | 966 (26.3%) |  |
| Gravidity |  |  |  |
| 1 | 44,750 (22.6%) | 1,125 (30.6%) | <0.001 |
| 2-4 | 85,627 (57.2%) | 2,041 (55.5%) |  |
| 5+ | 30,268 (20.2%) | 510 (13.9%) |  |
| Smoking status |  |  |  |
| No | 133,800 (89.4%) | 3,030 (82.4%) | <0.001 |
| Yes | 14,076 (9.4%) | 596 (16.2%) |  |
| Missing | 1,769 (1.2%) | 50 (1.4%) |  |
| Inflammatory bowel disease |  |  |  |
| No | 147,971 (98.9%) | 3,625 (98.6%) | 0.148 |
| Yes | 1,674 (1.1%) | 51 (1.4%) |  |
| Asthma |  |  |  |
| No | 114,827 (76.7%) | 2,685 (73.0%) | <0.001 |
| Yes | 34,818 (23.3%) | 992 (27.0%) |  |
| Diabetes mellitus |  |  |  |
| No | 146,168 (97.7%) | 3,521 (95.8%) | <0.001 |
| Yes | 3,477 (2.3%) | 155 (4.2%) |  |
| Gestational diabetes |  |  |  |
| No | 139,262 (93.1%) | 3,234 (88.0%) | <0.001 |
| Yes | 10,383 (6.9%) | 442 (12.0%) |  |
| Oligohydramnios |  |  |  |
| No | 145,356 (97.1%) | 3,517 (95.7%) | <0.001 |
| Yes | 4,289 (2.9%) | 159 (4.3%) |  |
| Antibiotic treatment during pregnancy |  |  |  |
| No | 96,173 (64.3%) | 2,314 (62.9%) | 0.103 |
| Yes | 53,472 (35.7%) | 1,362 (37.1%) |  |
| Pregnancy with history of pre-term labor |  |  |  |
| No | 143,027 (95.6%) | 3,558 (96.8%) | <0.001 |
| Yes | 6,618 (4.4%) | 118 (3.2%) |  |
| Decreased fetal movements |  |  |  |
| No | 146,931 (98.2%) | 3,575 (97.3%) | <0.001 |
| Yes | 2,714 (1.8%) | 101 (2.7%) |  |
| Delivery type |  |  |  |
| Vaginal birth | 119,200 (79.7%) | 2,528 (68.8%) | <0.001 |
| Assisted vaginal delivery | 5,869 (3.9%) | 214 (5.8%) |  |
| Cesarean section | 24,576 (16.4%) | 934 (25.4%) |  |
| **Infant** |  |  |  |
| Gestational age at birth, wks |  |  |  |
| Preterm < 37 | 6,780 (4.5%) | 223 (6.1%) | <0.001 |
| Term ≥37 | 142,865 (95.5%) | 3,453 (93.9%) |  |
| Gender |  |  |  |
| Female | 73,866 (49.4%) | 804 (21.9%) | <0.001 |
| Male | 75,779 (50.6%) | 2,872 (78.1%) |  |
| Birth weight, g |  |  |  |
| Mean (SD) | 3,240 (458) | 3,220 (498) | 0.063 |
| Median [Min, Max] | 3,250 [550, 5,850] | 3,240 [930, 5,520] |  |

Kaplan-Meier plots of the cumulative incidence of an ASD diagnosis for the offspring of mothers who received an influenza vaccination during pregnancy compared to those who did not are presented in **Fig. 2**. The cumulative incidence of ASD was significantly higher for infants born to mothers who received an influenza vaccination during pregnancy compared to unvaccinated mothers (0.036, 95%CI=0.034-0.039 vs. 0.029, 95%CI=0.028-0.030, **Fig. 2A**),with the highest cumulative incidence being found for the offspring of mothers vaccinated during the first trimester of pregnancy, followed by the third and second trimesters (**Fig. 2B**).

**Fig. 2.**
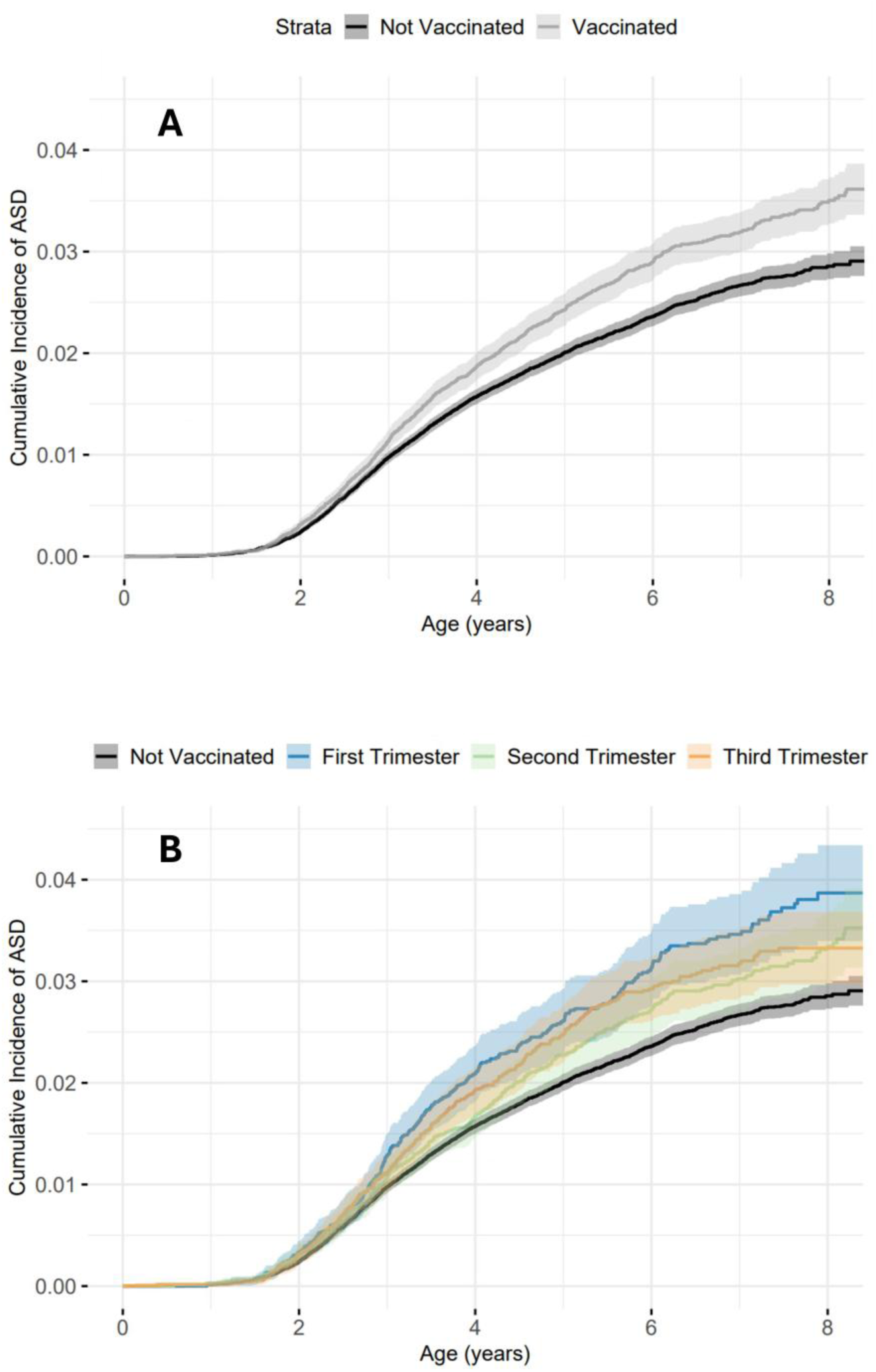
Kaplan-Meier Survival Plots of the Cumulative Incidence of ASD in Relation to Influenza Vaccine Exposure During Pregnancy. (A) Comparison of the cumulative incidence of ASD between mothers exposed (gray line) and unexposed (black line) to influenza vaccine during pregnancy. (B) Comparison of the cumulative incidence of ASD in the exposed group, stratified by pregnancy trimester exposure (blue, green, and orange lines representing the first, second, and third trimesters respectively), and the unexposed group (black line). Shaded areas denote the 95% confidence intervals for each exposure subgroup.

Cox regression analysis was conducted to explore the independent association between maternal influenza vaccination during gestation and a subsequent ASD diagnosis in the offspring (**Table 3**). Three regression models were built, starting with a univariate analysis that included only the exposure variable (null model); sociodemographic (Model I) and gestational (Model II) variables were then added aggregately to the regression analysis. In the univariate analysis, maternal influenza vaccination was significantly associated with a 1.22 increased risk of ASD in the offspring compared to no vaccination (HR=1.22, 95%CI=1.14-1.31), irrespective of the pregnancy trimester of vaccination (**Table 3**). However, adding sociodemographic and gestational variables to the regression model rendered the hazard ratio for the association of ASD with maternal influenza vaccination during gestation not significant (aHR = 0.97, 95%CI = 0.91- 1.05), suggesting that the observed association in the null model was due to confounding by the sociodemographic and gestational differences between the exposed and unexposed groups.

**Table 3.** Hazard ratios of ASD associated with influenza vaccine administration during pregnancy

|  | Total number of pregnancies | ASD cases (%) | Unadjusted hazard ratio (95%CI) | Model I<sup>a</sup> (95%CI) | Model II<sup>b</sup> (95%CI) |
| --- | --- | --- | --- | --- | --- |
| Unexposed during pregnancy | 113,960 | 2,578 (2.3) | - | - | - |
| Vaccination during pregnancy | 39,361 | 1,098 (2.8) | 1.22 (1.14-1.31) | 1.02 (0.95-1.10) | 0.97 (0.91-1.05) |
| Vaccination during first trimester | 10,489 | 316 (3.0) | 1.34 (1.19-1.50) | 1.09 (0.97-1.22) | 1.02 (0.91-1.15) |
| Vaccination during second trimester | 15,260 | 414 (2.7) | 1.15 (1.04-1.28) | 0.96 (0.86-1.07) | 0.92 (0.83-1.03) |
| Vaccination during third trimester | 13,612 | 368 (2.7) | 1.2 (1.09-1.35) | 1.04 (0.93-1.16) | 0.99 (0.89-1.12) |

<sup>a</sup>Adjusted for maternal age at delivery, district of residence, ethnicity.
<sup>b</sup>Adjusted for maternal age at delivery, maternal BMI, gravidity, smoking status, ethnicity, district of residence, maternal comorbidity (asthma, diabetes mellitus), gestational diabetes, oligohydramnios, pregnancy with history of pre-term birth, delivery type, gestational age at birth, infant gender, birth weight

Finally, we further examined the influence of maternal ethnicity and place of residence on our findings by applying the fully adjusted regression model to Bedouin Arabs living in Southern Israel and the other women in the study. These groups were characterized by maternal ethnicity and place of residence, two variables demonstrating remarkable difference in both maternal vaccination and ASD diagnosis rates (**Tables 1&2**). No significant association between maternal vaccination and ASD risk was seen in any of these subgroups (**Table S1**), suggesting no mediation effect of these sociodemographic variables on the association between maternal vaccination and ASD risk.

## Discussion

This study is among the largest and most comprehensive studies to investigate the association of influenza vaccination during pregnancy with the risk of ASD in the offspring. Our regression analyses show that influenza vaccination during pregnancy, regardless of the trimester in which the vaccination was administered, is not associated with ASD risk in the offspring. These findings largely align with the results reported in two other large cohort studies, one conducted in the U.S.(12) and the other in Sweden(13) The consistency of the results across these three large cohort studies involving different populations, with different sociodemographic and clinical characteristics, further reinforces the lack of effect of influenza vaccination during pregnancy on the risk of ASD in the offspring.

The finding of a higher ASD incidence in the offspring of vaccinated mothers compared to those of unvaccinated mothers can probably be attributed to confounding by sociodemographic and clinical characteristics, as suggested by our regression analyses. This confounding effect is likely due to the substantially lower proportion in the exposed group of Bedouin Arab women living in southern Israel. In Israel, the Bedouin subpopulation is known to differ from the general population in terms of socioeconomic status, health behaviors, and healthcare utilization (including adherence to vaccination).(17,18) For this subpopulation, there is also a lower rate of ASD diagnosis than in the general Israeli population.(19,20) Indeed, applying our regression analysis separately to Bedouin women and to other women in our study (**Table S1**) showed that in both groups, maternal influenza vaccination was not significantly associated with ASD risk in the offspring, despite the substantial differences between the groups both in rates of maternal influenza vaccination and in ASD prevalence in the offspring. These results further support the confounding effect of this ethnic difference in our study.

Our trimester-specific analysis revealed minor, non-significant differences in ASD risk associated with maternal influenza vaccination at different stages of pregnancy, with the highest risk observed in those vaccinated during the first trimester, followed by the third and second trimesters. These findings are consistent with those observed in the U.S. study(12), which also reported a higher ASD risk among offspring of mothers vaccinated during the first trimester of pregnancy. Nevertheless, unlike our study, the association of ASD with first-trimester vaccination in the U.S. study remained statistically significant even after adjustment for potential confounders.(12) The higher rate of ASD in offspring of mothers vaccinated against influenza during the first trimester compared to those vaccinated later in pregnancy in both studies may stem from the unique characteristics and health behaviors of women who favor being vaccinated early in pregnancy. These women may have greater health awareness and better access to medical care, two factors that could also increase the likelihood of an ASD diagnosis in their children.(21,22)

## Strengths and Limitations

This study features several significant strengths. In particular, the large sample size and extended follow-up period provided substantial statistical power and allowed for the accurate capture of ASD diagnoses over time. The median follow-up period of 5.9 years ensured a robust timeframe for analyzing long-term outcomes, further enhancing the reliability of the conclusions.

Additionally, the study included an analysis of stratification by trimester of exposure, enabling a nuanced understanding of the potential impact of the timing of maternal influenza vaccination on ASD risk. Moreover, the study actively sought to neutralize the influence of known genetic syndromes by excluding such cases from the analysis. These methodological strengths ensure that the findings contribute meaningful insights to the ongoing discourse on maternal vaccination and its implications for offspring health.

Importantly, this study also has some limitations that should be considered when interpreting results. First, the data utilized in the study was derived from a single healthcare organization (CHS), which may limit the generalizability of the findings to other populations in Israel.

However, CHS is the largest healthcare provider in the country, serving over 50% of the population, thereby making the sample highly representative. In addition, despite extensive efforts to adjust for relevant confounders, residual confounding from unmeasured factors cannot be entirely ruled out, as the etiology of ASD involves a multifactorial interplay of genetic, environmental, and prenatal influences. Despite familial or genetic risk factors being important contributors to ASD, this information was not available in the CHS database, limiting our ability to account for their potential effect on our results. Nonetheless, children with co-existing genetic syndromes relevant to ASD (Fragile X, Down syndrome) were excluded to help reduce potential confounding. In addition, parental education and household income were also not available in the CHS dataset. Nonetheless, designation of ethnicity may serve as an appropriate proxy for such socio-economic status in this region.(23,24) Finally, another possible limitation may be the lack of prospective diagnosis of ASD in the study. However, ASD diagnoses in Israel are typically determined by specialized clinicians (developmental pediatricians, neurologists, or psychiatrists) following DSM-5 criteria, with completion of formal diagnostic assessments allowing for high specificity of ascertainment within the CHS database.(25)

## Conclusion

These findings suggest that maternal influenza vaccination during pregnancy is not associated with the risk of ASD diagnosis in the offspring, regardless of the timing of vaccination. Thus, it addresses public concerns regarding the long-term safety of maternal influenza vaccination and provides reassurance for public health policies promoting influenza vaccination during pregnancy.

## Data Availability

All summary data produced in the present study are available upon reasonable request to the authors

## Acknowledgment

We thank Mrs. Inez Mureinik and Mrs. Leena Elbedour for critically reviewing and editing the manuscript.

Shachar Neeman and Idan Menashe, PhD, have full access to all the data in the study and take responsibility for the integrity of the data and the accuracy of the data analysis.

---

**Supplementary Table S1.** : Influenza vaccination and ASD risk in Bedouin Arab Women Living in the South of Israel vs. All Other Women in the Study

| Group | Exposure status | Total number of women | ASD cases (%) | aHR (95%CI)<sup>a</sup> |
| --- | --- | --- | --- | --- |
| Bedouin Arab women living in southern Israel | Unexposed | 31,496 | 187 (0.6%) | - |
|  | Exposed | 3,715 | 30 (0.8%) | 1.23 (0.82-1.83) |
| Other women in the study (non-Bedouin) | Unexposed | 82,464 | 2,391 (2.9%) | - |
|  | Exposed | 35,646 | 1,061 (3.0%) | 0.96 (0.90-1.04) |

<sup>a</sup>Adjusted for maternal age at delivery, maternal BMI, gravidity, smoking status, district of residence, maternal comorbidity (asthma, diabetes mellitus), gestational diabetes, oligohydramnios, pregnancy with history of pre-term birth, delivery type, gestational age at birth, infant gender, and birth weight

